# Estimating the impact of the *w*Mel release program on dengue and chikungunya incidence in Rio de Janeiro, Brazil

**DOI:** 10.1101/2022.03.29.22273035

**Authors:** Gabriel Ribeiro dos Santos, Betina Durovni, Valeria Saraceni, Thais Irene Souza Riback, Sofia B. Pinto, Katherine L. Anders, Luciano A. Moreira, Henrik Salje

## Abstract

**Background:** Introgression of the insect bacterium *Wolbachia* into *Aedes aegypti* mosquito populations been shown in randomised and non-randomised trials to reduce the incidence of dengue in treated communities, however evidence for the real-world effectiveness of large-scale *Wolbachia* mosquito deployments for arboviral disease control in endemic settings is still limited and no effectiveness studies have been conducted for chikungunya virus. A large *Wolbachia* (*w*Mel strain) program was implemented in 2017 in Rio de Janeiro, Brazil. Here we assess the impact of the release program on dengue and chikungunya incidence.

**Methods and findings:** The program released 67 million *w*Mel infected mosquitoes across 28,489 release locations over a 86.8km2 area in Rio de Janeiro between August 2017 and the end of 2019. Following releases, mosquitoes were trapped and the presence of *w*Mel determined. To assess the impact of the release program on dengue and chikungunya incidence, we used spatiotemporally explicit models applied to geocoded dengue (N=194,330) and chikungunya cases (N=58,364) from 2010 (2016 for chikungunya) to 2019 from across the city. On average, 32% of mosquitoes collected from the release zones between 1 and 29 months after releases were positive for *w*Mel. Reduced *w*Mel introgression occurred in locations and seasonal periods when dengue and chikungunya cases were historically high. Despite incomplete introgression, we found that the releases were associated with a 38% (95%CI: 32-44%) reduction in dengue incidence and a 10% (95%CI: 4-16%) reduction in chikungunya incidence.

**Conclusions:** Stable establishment of *w*Mel in this diverse, urban setting appears more complicated than has been observed elsewhere. However, even intermediate levels of *w*Mel appear to reduce the incidence of two different arboviruses.

## Introduction

Dengue virus continues to circulate endemically across global tropical and subtropical regions, causing an estimated 50 million symptomatic infections per year^1^. In addition, large-scale outbreaks of chikungunya virus, spread by the same *Aedes* mosquitoes, have become increasingly common. The use of *Wolbachia* (*w*Mel strain) infected mosquitoes is a promising new technology to reduce transmission^2^. *Wolbachia* is an intracellular bacterium that can be stably inserted into *Ae. aegypti*^*3*^. *w*Mel infected mosquitoes have been shown to have reduced ability to harbour and transmit different arboviruses, including dengue and chikungunya viruses^3–7^.

When released, *w*Mel-infected *Ae. aegypti* mate with the wildtype population. As the offspring from an infected female all have *w*Mel and offspring from an infected male and a wildtype female are non-viable, this drives the introgression of *w*Mel into the mosquito population. Results of field trials in multiple countries have shown successful establishment of *w*Mel in the local Aegypti population following a release period of several weeks or months^8–12^. In a cluster randomised controlled trial of *w*Mel-infected *Ae. aegypti* deployments in Indonesia, *w*Mel quickly reached greater than 90% penetration which was followed by a significant reduction in dengue incidence and dengue hospitalisations in intervention areas^9,13^. *w*Mel has also been shown to reduce both dengue and chikungunya incidence in Niteroi, Brazil^14^.

Except in extreme scenarios where case numbers crash to virtually zero, understanding the impact of spatially targeted interventions is complicated as dengue and chikungunya cases vary substantially over space and time, driven by local variations in immunity, human behaviours, mosquito density, population density, building constructions and climate among other factors^15–17^. Further, movement of people outside a release zone means that local residents can still become cases, even if the intervention is 100% effective. Finally, most infections are not detected, due to limited symptoms or lack of healthcare seeking^18,19^.

As *w*Mel is deployed at increasing scale, we need a robust understanding of the impact of these releases on arboviral disease incidence in a range of epidemiological settings. Here, we study the impact of *w*Mel releases in Rio de Janeiro, Brazil, on dengue and chikungunya case occurrence in that city. Entomological results one year after the conclusion of releases in the first two release areas in Rio showed successful *w*Mel introgression overall, but with heterogeneity in *w*Mel prevalence at the neighbourhood level (Gesto et al Frontiers in Micro 2021). We make use of the city of Rio’s detailed public health systems, where dengue and chikungunya cases from all hospitals and health clinics throughout the city have been systematically geocoded for 9 years. We develop spatially explicit mathematical models to fit the timing and location of cases and estimate the impact of *w*Mel on both dengue and chikungunya occurrences.

## Material and methods

### Setting and wMel field implementation

Rio de Janeiro is the second biggest city of Brazil with 6.7 million inhabitants over 1,260 km^2^. It is a patchwork of highly-dense flat urban areas and uninhabited mountains covered with tropical forest. The *w*Mel release program started in the Northwest of the city in August 2017^20^. The release area was subdivided into 5 zones (RJ1, RJ2, RJ3.1, RJ3.2 and RJ3.3) covering a total area of 86.8 km^2^ with around 890.000 inhabitants and releases phased through the different zones^21^.

A pilot test in the city identified that insecticide resistance was widespread in wildtype *Ae. aegypti*, which can hinder the successful establishment of *w*Mel in an area. The release program therefore crossed *w*Mel females mosquitoes with wildtype males, which allowed a better matching of genetic profiles between *w*Mel infected and field mosquitoes^22^. All releases used this newly developed locally-matched insecticide resistant mosquito strain. Releases started in August 2017 and continued until the end of 2019. Release points were distributed regularly every 50m in five release zones. Around 100 *w*Mel-infected mosquitoes were released each time. A network of 1,168 BG sentinel traps was used to monitor *w*Mel introgression. The traps were regularly distributed throughout the release area with an average distance between two adjacent traps of 250m. A trap was set four weeks after the first release in the area and collection was conducted every other week from then on (N=36,894 trap collections). Up to ten *Ae. aegypti* mosquitoes (male and female) per collection were tested individually for *w*Mel using qPCR. This gives an estimation of the global proportion of mosquitoes that were infected in a given trap at a given time.

### Case data

Every case in Rio de Janeiro that presents to a healthcare facility and that is suspected to be dengue or chikungunya must be recorded in the national surveillance system database (SINAN). Suspected dengue cases are defined as those that present with fever and at least two of the following manifestations: nausea/vomiting, rashes, myalgia/arthralgia, headache/retro-orbital pain, petechiae/positive tourniquet test, and leukocytopenia. Suspected chikungunya cases are defined as patients with fever and arthralgia or arthritis. A differential diagnosis between chikungunya is based on the duration of fever (up to 7 days for dengue and up to 3 days) and the intensity of arthralgia, which is more intense in chikungunya. Lymphocytopenia is also frequent in chikungunya cases whereas it is uncommon for dengue cases^23^. A subset of cases are laboratory confirmed. For dengue, the confirmation was conducted using PCR (12.0% of cases, N=33,425), whereas for chikungunya, confirmation is mainly conducted through IgM serology (25.2% of all suspected cases, N=14,528).

All dengue and chikungunya cases are systematically geolocated by the Rio de Janeiro city health department using home address information where possible. A subset of notified cases were not geolocatable, 63,598 cases for dengue (18% of total cases) and 4,710 cases for chikungunya (8% of total cases) and were therefore not included in the analysis.

### Spatial model

For our modelling approach we divided up the project area into 500 × 500 metre cells (N= 465 cells). We then counted the number of dengue and chikungunya cases occurring within each 30 day period in each cell from 2010-2019 (N=117 time periods, resulting in 54,405 total space-time units). We used WorldPop data to obtain detailed estimates of the distribution of the underlying population throughout the project area^24^.

We constructed Poisson regression models to separately fit the number of dengue cases and chikungunya cases for each space-time unit throughout the project area during the period 2010-2019 for dengue and during the period 2016-2019 for chikungunya. In order to incorporate the spatial correlation in the location of dengue and chikungunya cases, we used Integrated Nested Laplace Approximation, as implemented in R-INLA^25^. This approach allowed us to introduce a spatial correlation term that explicitly incorporates the spatial dependence between locations. In addition, to account for temporal correlation in the timing of cases, we used a temporally structured random effect by using an order one autoregressive model to the monthly time variable. We used the log of population size within the cell as an offset to the model.

To estimate the impact of the *w*Mel release program, we considered three separate measures. First, we used a binary variable where each space-unit was coded as 1 if *w*Mel was detected in *Ae. Aegypti* mosquitoes across the traps within that location and within that month. Second, we used the actual proportion of *Aedes* mosquitoes that were infected by *w*Mel across these traps, using non-overlapping bins (0%, 0.1-10%, 10.1-20%, 20.1-30%, 30.1-40%, 40.1-50%, 50.1-60%, >60%). Finally, we considered *w*Mel as a continuous variable and estimated the reduction in chikungunya/dengue incidence for each unit increase in *w*Mel. In all models, space-time units prior to the initiation of releases were given a value of 0. Space-time units with no traps and with zero catches were removed from the main analysis.

### Sensitivity analysis

To assess the robustness of our approach, we conducted a range of sensitivity analyses. To test whether the released *Ae. aegypti* were being re-captured in the traps (leading to falsely high estimates of *w*Mel) and potentially driving our estimates, we conducted a sensitivity analysis where we excluded space-time units that occurred within a short amount of time following releases. Two different exclusion criteria were designed. First, we excluded space-time units within which releases occurred, this resulted in the exclusion of 57% space-time units with a non-zero *w*Mel value. The second, more exclusive criterion was to exclude from the analysis space-time units both for the month of the release and the subsequent month. This approach resulted in 67% of space-time units with non-zero *w*Mel values being excluded.

In a further sensitivity analysis we repeated the analysis using an augmented dataset that inferred the number of *w*Mel-infected *Ae. aegypti* captured in places with missing %*w*Mel data. Missing %*w*Mel estimates occurred in places where either no traps were set up despite releases having already occurred in the area or when traps did not capture any mosquitoes. We used a separate spatial regression model to estimate the number of *w*Mel positive mosquitoes within each space-time unit. The model to estimate %*w*Mel can be summarised by *Ni*(*s, t*) | η(*s, t*) ∼*Poisson*(*Nt*(*s, t*) * (*u*(*s*) + *v*(*t*)), where *Ni*(*s, t*) is the number of infected *Ae. aegypti* captured in a given cell, *Nt*(*s, t*) is the total number of *Ae. aegypti* aptured in the same cell, *u*(*s*) is a spatially structured random effect and *v*(*t*) is a temporally structured random effect. Both random effects were defined in the same way as for the case-count model (Figure S1). We fit the model using the observed mosquito count and %*w*Mel data. We then replaced the space-time locations with missing %*w*Mel estimates by values predicted by this *w*Mel model leading to an increase of the dataset by 23%. We also ran a sensitivity analysis where all the *w*Mel data set was entirely replaced by values predicted by the *w*Mel prediction model (and not just in the space-time locations with missing %*w*Me estimates).

### Model fit

We examined model fit by comparing predicted case counts within space-time units to that actually observed. We split the dataset into training and testing sets. We then fit the model using the training set only and predicted the number of cases per space-time unit in the testing set. We used two different ways to split the case dataset into testing and training datasets. The first way was to randomly split the case dataset into two equal parts, with each space-time unit having an equal probability of being within each set. The second way was to split up large spatiotemporal regions, consisting of 20% of the global dataset, as the testing dataset. These regions had an average size of 5 km ∧2 and a temporal length of a year. This means that the model was asked to predict the count number in a location it has no information on through a whole year.

### Ethical considerations

This study was approved by the Brazilian National Institutional Review Board (CONEP - 59175616.2.0000.0008).

## Results

Between August 2017 and December 2019 an estimated 67 million *w*Mel-infected mosquitoes were released in 658,179 individual release events at 28,489 different locations in the five release zones (Figure 1A-B). There were an average of 24,377 releases per month. There were a total of 36,894 mosquito trap collection events of which 23,071 contained at least one *Ae. aegypti* mosquito. We found that the overall proportion of trapped mosquitoes that were *w*Mel positive was 32% (Figure 1C), although there were differences across the release region and over time, with higher prevalence observed in release zone RJ1, where an average of 52.3% was observed and lower values in the latter release zone RJ3.1, where the average was 19.8% (Figure 1B).

**Figure 1.**
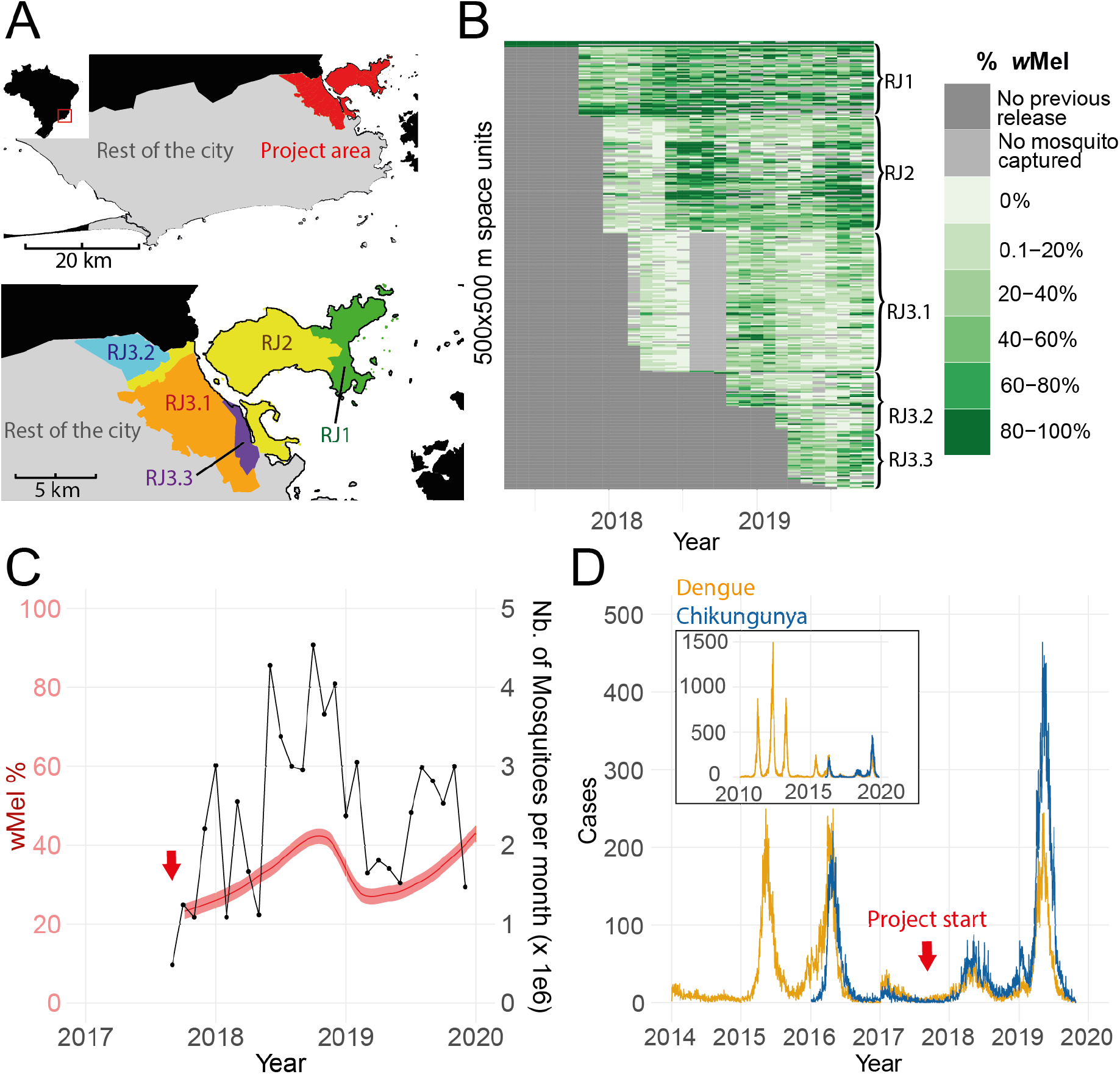
Details on *w*Mel release program and dengue and chikungunya incidence in Rio de Janeiro. **(A)** Map of Rio de Janeiro with a detail on the project area, showing the different project subareas. Inset shows Rio within Brazil. **(B)** Proportion of *w*Mel infected *Ae. aegypti* found in traps for each spatiotemporal cell. **(C)** Number of mosquitoes released and the level of introgression. Black broken line represents the total amount of mosquitoes released per month. Red ribbon represents the proportion of *Ae. aegypti* captured in traps that were infected by *w*Mel. **(D)** Temporal distribution of dengue cases (orange) and chikungunya (blue) cases identified Rio de Janeiro, including both the project area and the rest of the city. Inset shows cases recorded since 2010.

Between 2010 and 2019 there were 283,270 cases of dengue reported in Rio’s healthcare centres with an average of 31,474 cases per year. Chikungunya became a notifiable disease in 2016 and has since been continuously reported with an average of 14,426 cases per year (57,705 total chikungunya cases since it first emerged) (Figure 1D). Both chikungunya and dengue cases were found throughout the city (Figure S2).

We observed strong seasonality in the case data and the mosquito data. For both dengue and chikungunya, we found that transmission peaks during March-April (Figure 2A). The number of *Ae. aegypti* and *Ae. albopictus* trapped as part of the study also followed a similar seasonal trend, peaking in March (Figure 2B). By contrast, the proportion of mosquitoes that were infected with *w*Mel was inversely correlated with the number of cases (rho = -0.82), dropping to around 25% in March and April and peaking at 49% in August, when disease incidence is lowest (Figure 2C). The correlation of *w*Mel with the number of releases by month was less substantial (rho= -0.34).

**Figure 2.**
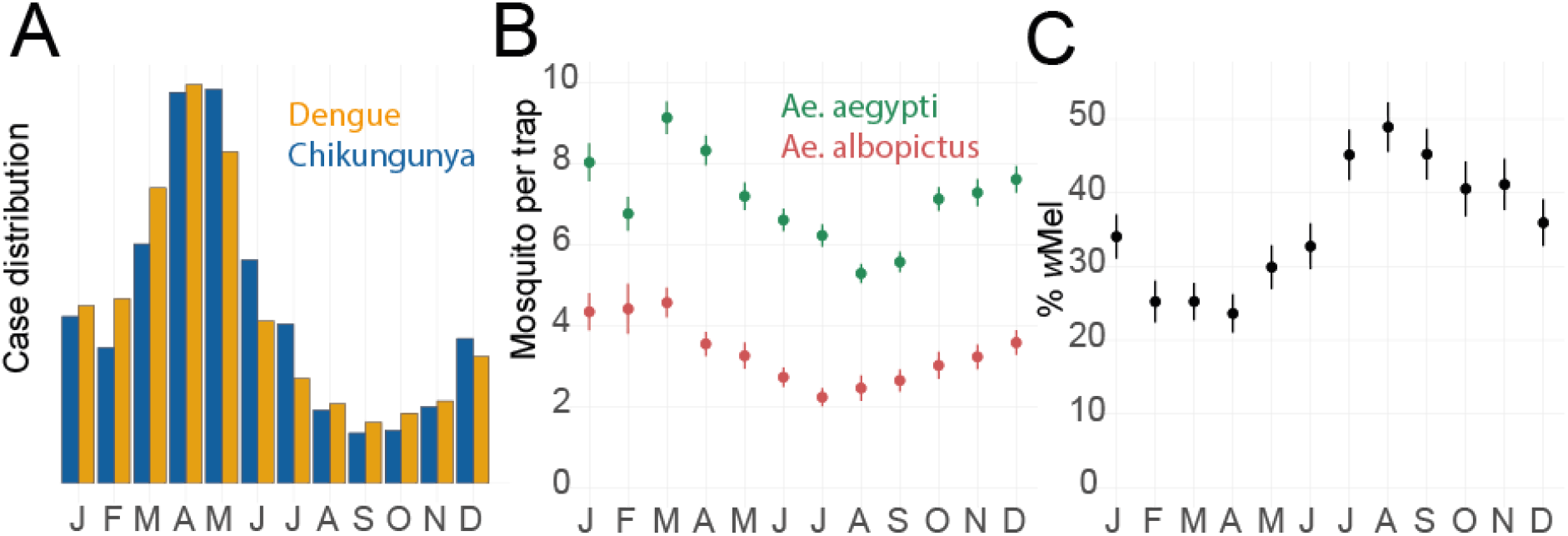
Seasonal pattern of cases and mosquito levels. **(A)** Monthly distribution of dengue (2010-2019) and chikungunya cases (2016-2019). **(B)** Monthly distribution of *Aedes aegypti* and *Aedes albopictus* mosquitoes found in traps (2017-2019). **(C)** Average proportion of trapped female *Aedes aegypti* mosquitoes that carried *w*Mel by month across the project area (2017-2019).

To explore the overall relationship between *w*Mel introgression and both historic (pre release program) and subsequent case incidence, we initially compared the *w*Mel prevalence in each month-space unit with the number of cases reported in the same cell and time period across different years (Figure 3). We looked at case incidence at the same time period as the *w*Mel data (solid lines) as well as incidence in the equivalent time period in previous years (dashed lines). This exercise demonstrated that *w*Mel introgression was more successful in months and locations where dengue and chikungunya case incidence tended to be lower each year.

**Figure 3.**
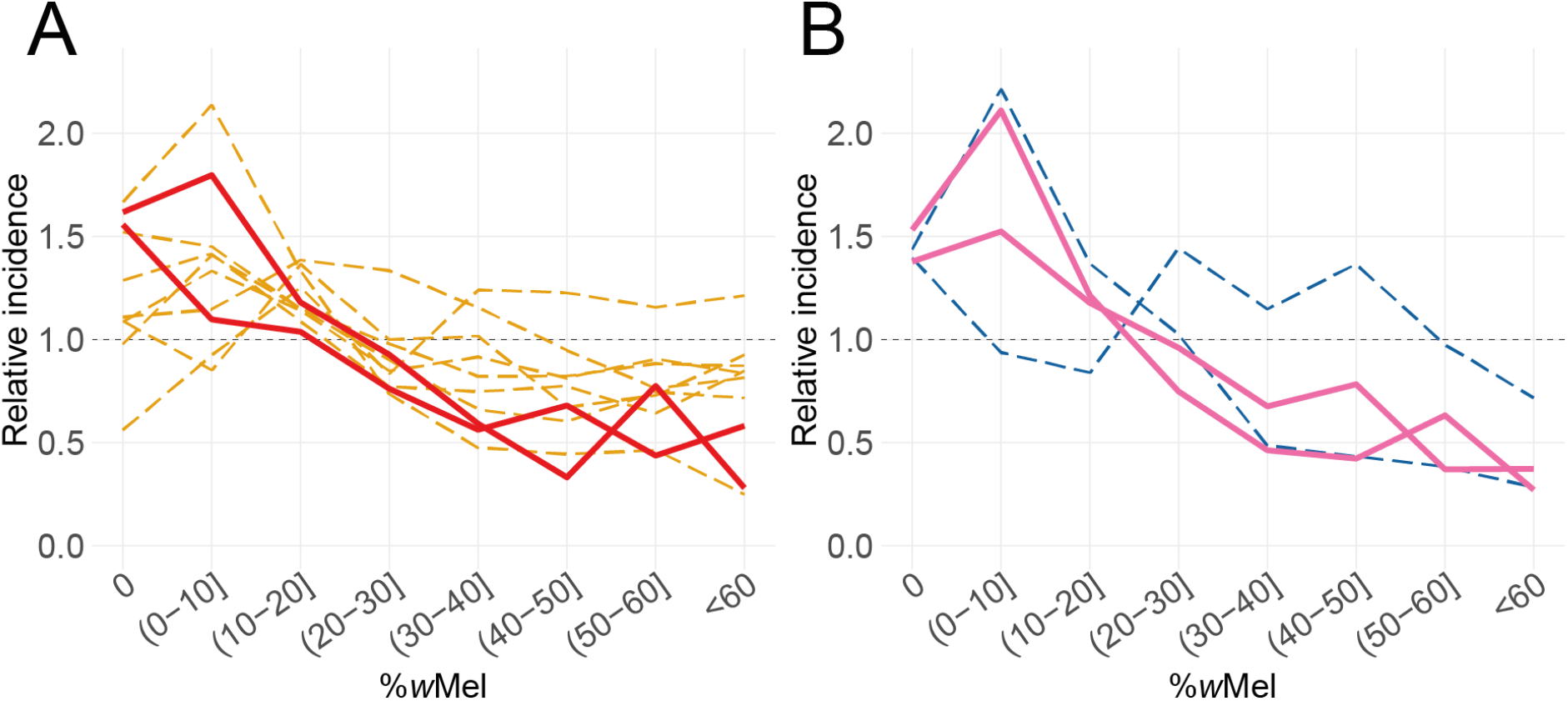
Introgression success as a function of historical and future case incidence. Comparison of the average monthly introgression of *w*Mel within a 500m x 500m cell and the standardised dengue **(A)** and chikungunya **(B)** incidence in that location and month. Each line represents the incidence from a different year. Dashed lines are years before the start of the release program (2010-2017 for dengue 2016-2017 for chikungunya). Solid lines represent the years 2018 and 2019, which are after the release program started. We standardised the dengue and chikungunya incidence by dividing by the overall mean incidence in that year. This allows us to compare large outbreak years with smaller years on the same plot.

To estimate the impact of the intervention accounting for underlying heterogeneities in space and time in both the case data and the mosquito data, we fit a spatially explicit model to where cases were found. 2209 space-time units (34% of the project area) had no trapped *Ae. aegypti* mosquitoes and were excluded from the main analysis. We found the presence of *w*Mel in local *Ae. aegytpi* had a strong effect on the number of cases in that location at that time (Figure 4). On average, the dengue incidence within a space-time unit where *w*Mel was detected was 0.62 (95%CI: 0.56-0.68) times the incidence in space-time units without any *w*Mel detected. The relative incidence of chikungunya was 0.90 (95%CI: 0.84-0.96) (Figure 4A). The model estimated that spatial correlation between the location of dengue cases within any month extended to 1460m (95% CI: 1071-1815) and to 2081m (95% CI: 1594-2610) for chikungunya cases.

**Figure 4.**
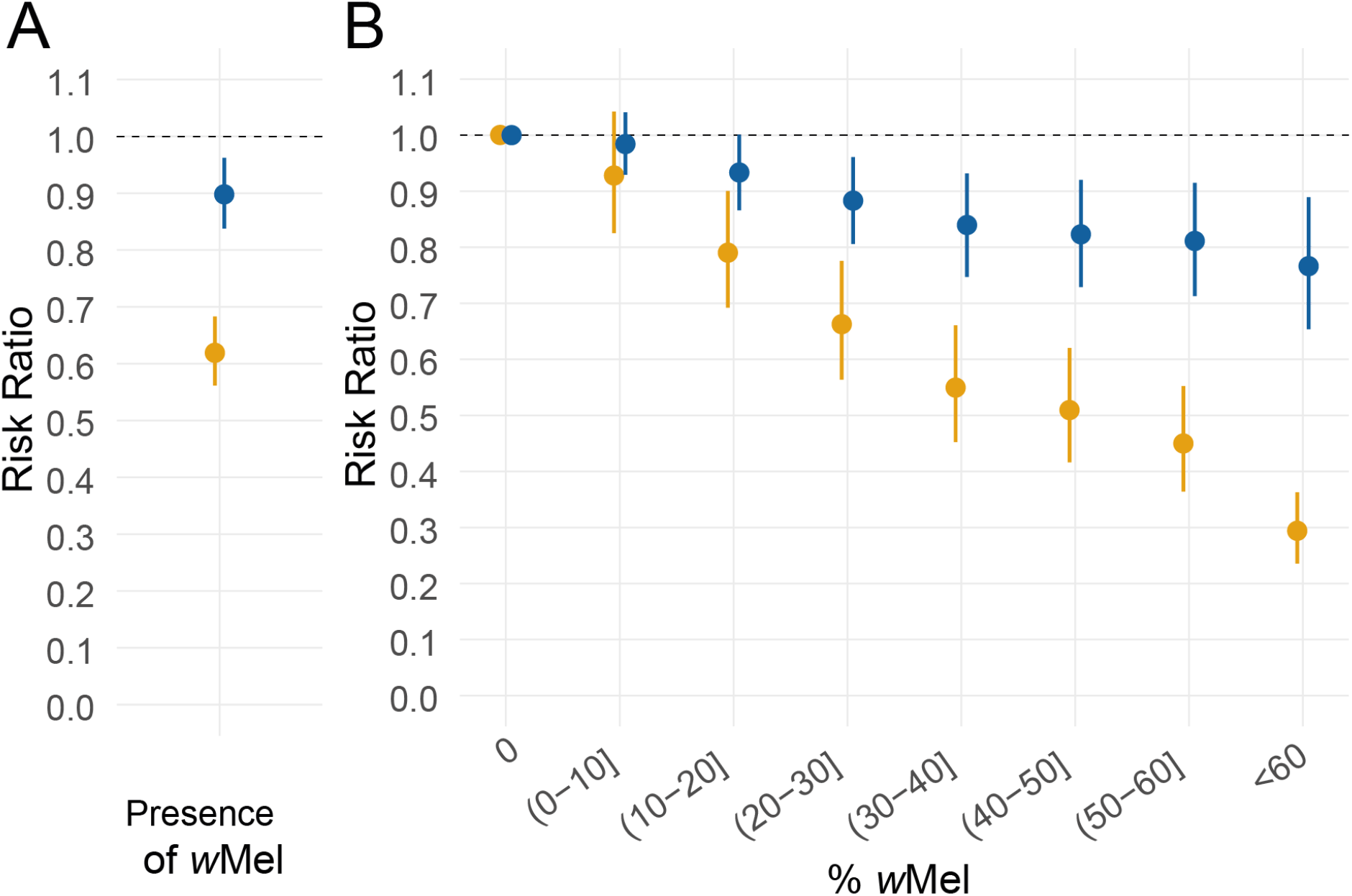
Results of spatio-temporal models. **(A)** Estimated overall relative incidence of dengue (orange) and chikungunya (blue) in locations and time periods where *w*Mel presence was recorded as compared to where there was no *w*Mel. **(B)** Relative incidence of dengue (orange) and chikungunya (blue) in locations and time periods as a function of the proportion of *Ae. aegypti* mosquitoes that had *w*Mel. Space-time units within the study area with no detected *w*Mel are the reference.

We next fit separate models that explored the relationship between the observed proportion of *Ae. aegypti* mosquitoes within each cell and month that were infected with *w*Mel and the dengue and chikungunya incidence in that space-time unit. We found that where 0.1-10% of the mosquitoes had *w*Mel there was 0.93 (95% CI: 0.63-0.79) times the incidence of dengue and 0.98 (95% CI: 0.93-1.04) the incidence of chikungunya as locations with no *w*Mel in that area (Figure 4B). This fell to 0.29 (95% CI: 0.24-0.36) for areas with more than 60%*w*Mel prevalence for dengue and 0.77 (95% CI: 0.65-0.89) for chikungunya. In separate models that considered wMel as a continuous variable, we estimated that each 10% increase in wMel was associated with 0.85 times the incidence of dengue (95%CI: 0.83-0.87) and 0.96 times the incidence of chikungunya (95%CI: 0.94-0.97) (Figure S3).

To ensure our results were not driven by the recapture of recently released mosquitoes, we repeated our analysis on a dataset where all locations where a release event had occurred within a previous month had been removed. As traps were not placed in all locations at all times leading to missing estimates of %*w*Mel in some space-time units, we also conducted a separate sensitivity analysis where we initially predicted the %*w*Mel in all space-time locations of the study region (Figure S1). In both these sensitivity analyses, we observed a consistent pattern of the impact of *w*Mel on both dengue and chikungunya incidence (Figure S4).

In order to assess the performance of our model we repeatedly removed randomly selected space-time locations from the model (held out locations) and predicted the number of cases in those locations using model fit on the remaining data. We then compared our estimates with the observed number of cases in that location. We found good correlation between the predicted and observed number of cases (Figure S5A), including when we held-out 50% of randomly selected cell-month units (Figure S5B), and when we held out randomly selected spatially clustered cells (5km2 in area) for one year at a time (Figure S5C).

## Discussion

We have critically assessed the impact of a *w*Mel release program on dengue and chikungunya incidence in a diverse, urban setting. By December 2019, 29 months after the commencement of phased releases, *w*Mel prevalence in local *Ae. aegypti* was between 27% and 60% in the five release areas in Rio de Janeiro^8,12,26^. Using a spatially and temporally explicit modelling framework we have demonstrated that despite heterogeneous *w*Mel prevalence, *w*Mel releases still resulted in lower incidence of both dengue and chikungunya viruses during the first two years post-intervention, highlighting the potential of this technology.

It has not previously been possible to quantify the protective effect of low-moderate wMel prevalence, because rapid establishment has generally been observed following *w*Mel deployments in other settings^8,9,12^and arboviral case notification data is not commonly available at a high spatial resolution^14^. Here we report a dose-response relationship between *w*Mel levels and relative reductions in dengue and chikungunya case incidence. A small but significant protective effect against dengue was seen even at very low local *w*Mel prevalence (≤10%) and where *w*Mel was >60% the protective effect was 76% (95%CI 64-71%), which is comparable to results reported previously (using different methods) from the neighbouring municipality of Niteroi, and from Indonesia^9^.

The reasons why *w*Mel has been unable to become quickly established in Rio de Janeiro despite large numbers of releases are unclear. Underlying dengue and chikungunya incidence in the city is highly heterogeneous. We found that *w*Mel introgression was lower in areas that annually see high disease incidence. There were also seasonal fluctuations in *w*Mel introgression with lower levels observed in February to May, which represent the hottest periods of the year. High temperatures have been linked to lower *w*Mel acquisition in laboratory studies ^27^. The release program also made fewer releases during the summer months, which may contribute to this observed seasonal effect. The areas of the city with stubbornly high dengue and chikungunya incidence may have factors that also complicate the *w*Mel release program, including large, heterogeneously distributed baseline mosquito populations, or are in areas that are hard to access, including favela communities. *Ae. albopictus* also circulates in Rio but has not been incriminated as being involved in dengue and chikungunya incidence ^28^. A role for *Ae. albopictus* in affecting *w*Mel introgression in *Ae. aegypti* remains unclear.

Cluster randomised trials, such as the one conducted in Yogyakarta and also currently being conducted in Belo Horizonte^29^, Brazil (Trial ID: NCT04514107) provide a gold standard measure of the effectiveness of *w*Mel on disease incidence. However, these large trials may not always be feasible, especially when resources are limited. Our study highlights how the systematic geocoding of cases provides a valuable resource to understand where incidence is concentrated that can also act as a reference point to evaluate the impact of spatially targeted interventions. However, we have shown that alongside detailed data, we need to use structured models to appropriately measure these datasets. For example, our finding that *w*Mel introgression achieved in a location was correlated with the incidence of dengue and chikungunya in that same location in years prior to the intervention, highlighted the complexity of using observational case data to understand the impact of an intervention whose penetration is itself spatially and temporally uneven. It is only through spatiotemporally structured models that we can disentangle these different correlation structures to identify the underlying impact of the intervention.

Our study considers data to the end of 2019. Increasing the period of analysis with additional years would provide important insight into the durability of the intervention. Unfortunately, the COVID-19 situation has had a significant impact on the mosquito release program and the city health department. In particular, the systematic geocoding of dengue and chikungunya cases by the city health department largely stopped in 2020. By the end of 2021, additional ovitrapping data showed that the average introgression level across the project area was at over 50%, suggesting complete introgression may yet be possible. The study also has other limitations. In particular, individuals move around the city and arboviral cases are notified based on place of residence but may have acquired their infection elsewhere. Further, the majority of the cases are based on clinical presentation alone, and an unknown proportion will have a non-dengue cause of febrile illness. There were insufficient numbers of confirmed cases within the study area to rely on confirmed cases only in our analyses. In the event that the misdiagnosed cases were caused by other pathogens, such as influenza that would not be affected by the mosquito release program, our estimates of the impact of *w*Mel would be biassed towards the null and the true impact may be larger.

*w*Mel continues to appear to be a promising technology with a significant impact of reducing the public health burden from different arboviruses within the same community. A major challenge is achieving establishment in complex urban communities such as Rio. Understanding why the introgression of *w*Mel into *Ae. aegypti* populations are more rapid and homogeneous in some locations than in others will help underpin its future success. The analytical framework we present can be applied to evaluate the effectiveness of other spatially and temporally targeted interventions.

## Data Availability

All data produced in the present study are available upon reasonable request to the authors

## References

1 Bhatt S, Gething PW, Brady OJ, et al. The global distribution and burden of dengue. Nature 2013; 496: 504–7.

2 Hoffmann AA, Montgomery BL, Popovici J, et al. Successful establishment of Wolbachia in Aedes populations to suppress dengue transmission. Nature 2011; 476: 454–9.

3 Walker T, Johnson PH, Moreira LA, et al. The wMel Wolbachia strain blocks dengue and invades caged Aedes aegypti populations. Nature 2011; 476: 450–3.

4 Pereira TN, Rocha MN, Sucupira PHF, Carvalho FD, Moreira LA. Wolbachia significantly impacts the vector competence of Aedes aegypti for Mayaro virus. Sci Rep 2018; 8: 1–9.

5 Dutra HLC, Rocha MN, Dias FBS, Mansur SB, Caragata EP, Moreira LA. Wolbachia Blocks Currently Circulating Zika Virus Isolates in Brazilian Aedes aegypti Mosquitoes. Cell Host Microbe 2016; 19: 771–4.

6 van den Hurk AF, Hall-Mendelin S, Pyke AT, et al. Impact of Wolbachia on Infection with Chikungunya and Yellow Fever Viruses in the Mosquito Vector Aedes aegypti. PLoS Negl Trop Dis 2012; 6. DOI:10.1371/journal.pntd.0001892.

7 Ye YH, Carrasco AM, Frentiu FD, et al. Wolbachia reduces the transmission potential of dengue-infected Aedes aegypti. PLoS Negl Trop Dis 2015; 9: 1–19.

8 O’Neill SL, Ryan PA, Turley AP, et al. Scaled deployment of Wolbachia to protect the community from dengue and other aedes transmitted arboviruses. Gates Open Research 2018; 2: 1–27.

9 Indriani C, Tantowijoyo W, Rancès E, et al. Reduced dengue incidence following deployments of Wolbachia -infected Aedes aegypti in Yogyakarta, Indonesia : a quasi-experimental trial using controlled interrupted time series analysis. 2020.

10 Hoffmann AA, Iturbe-Ormaetxe I, Callahan AG, et al. Stability of the wMel Wolbachia Infection following Invasion into Aedes aegypti Populations. PLoS Negl Trop Dis 2014; 8. DOI:10.1371/journal.pntd.0003115.

11 Schmidt TL, Barton NH, Rašić G, et al. Local introduction and heterogeneous spatial spread of dengue-suppressing Wolbachia through an urban population of Aedes aegypti. PLoS Biol 2017; 15: 1–28.

12 Ryan PA, Turley AP, Wilson G, et al. Establishment of wMel Wolbachia in Aedes aegypti mosquitoes and reduction of local dengue transmission in Cairns and surrounding locations in northern Queensland, Australia. Gates Open Research 2020; 3: 1547.

13 Utarini A, Indriani C, Ahmad RA, et al. Efficacy of Wolbachia-infected mosquito deployments for the control of dengue. N Engl J Med 2021; 384: 2177–86.

14 Pinto SB, Riback TIS, Sylvestre G, et al. Effectiveness of Wolbachia-infected mosquito deployments in reducing the incidence of dengue and other Aedes-borne diseases in Niterói, Brazil: a quasi-experimental study. DOI:10.1101/2021.01.31.21250726.

15 Salje H, Lessler J, Endy TP, et al. Revealing the microscale spatial signature of dengue transmission and immunity in an urban population. Proc Natl Acad Sci U S A 2012; 109: 9535–8.

16 Salje H, Lessler J, Paul KK, et al. How social structures, space, and behaviors shape the spread of infectious diseases using chikungunya as a case study. Proc Natl Acad Sci U S A 2016; 113: 13420–5.

17 Bavia L, Melanda FN, de Arruda TB, et al. Epidemiological study on dengue in southern Brazil under the perspective of climate and poverty. Sci Rep 2020; 10: 2127.

18 Stanaway JD, Shepard DS, Undurraga EA, et al. The global burden of dengue: an analysis from the Global Burden of Disease Study 2013. Lancet Infect Dis 2016; 16: 712–23.

19 Salje H, Cummings DAT, Rodriguez-Barraquer I, et al. Reconstruction of antibody dynamics and infection histories to evaluate dengue risk. Nature 2018; 557: 719–23.

20 Gesto JSM, Pinto S, Dias FBS, et al. Large-scale deployment and establishment of Wolbachia into the Aedes aegypti population in Rio de Janeiro, Brazil. DOI:10.1101/2021.04.29.441982.

21 Durovni B, Saraceni V, Eppinghaus A, et al. The impact of large-scale deployment of Wolbachia mosquitoes on arboviral disease incidence in Rio de Janeiro and Niterói, Brazil : study protocol for a controlled interrupted time series analysis using routine disease surveillance data [ version 1 ; pee. 2019; : 1–11.

22 Garcia G de A, Sylvestre G, Aguiar R, et al. Matching the genetics of released and local Aedes aegypti populations is critical to assure Wolbachia invasion. PLoS Negl Trop Dis 2019; 13: e0007023.

23 [No title]. https://bvsms.saude.gov.br/bvs/publicacoes/guia_vigilancia_saude_3ed.pdf (accessed Dec 7, 2020).

24 Worldpop. WorldPop. http://www.worldpop.org (accessed Aug 24, 2020).

25 Lindgren F, Rue H. Bayesian Spatial Modelling with R - INLA. J Stat Softw 2015; 63. DOI:10.18637/jss.v063.i19.

26 Indriani C, Tantowijoyo W, Rancès E, et al. Reduced dengue incidence following deployments of Wolbachia-infected Aedes aegypti in Yogyakarta, Indonesia: a quasi-experimental trial using controlled interrupted time series analysis. Gates Open Research 2020; 4: 50.

27 Ulrich JN, Beier JC, Devine GJ, Hugo LE. Heat Sensitivity of wMel Wolbachia during Aedes aegypti Development. PLoS Negl Trop Dis 2016; 10: e0004873.

28 Parra MCP, Lorenz C, de Aguiar Milhim BHG, et al. Detection of Zika RNA virus in Aedes aegypti and Aedes albopictus mosquitoes, São Paulo, Brazil. Infect Genet Evol 2022; 98: 105226.

29 Collins MH, Potter GE, Hitchings MDT, et al. EVITA Dengue: a cluster-randomized controlled trial to EValuate the efficacy of Wolbachia-InfecTed Aedes aegypti mosquitoes in reducing the incidence of Arboviral infection in Brazil. Trials. 2022; 23. DOI:10.1186/s13063-022-05997-4.

