## supplementary_figures for "Estimating the impact of the *w*Mel release program on dengue and chikungunya incidence in Rio de Janeiro, Brazil"

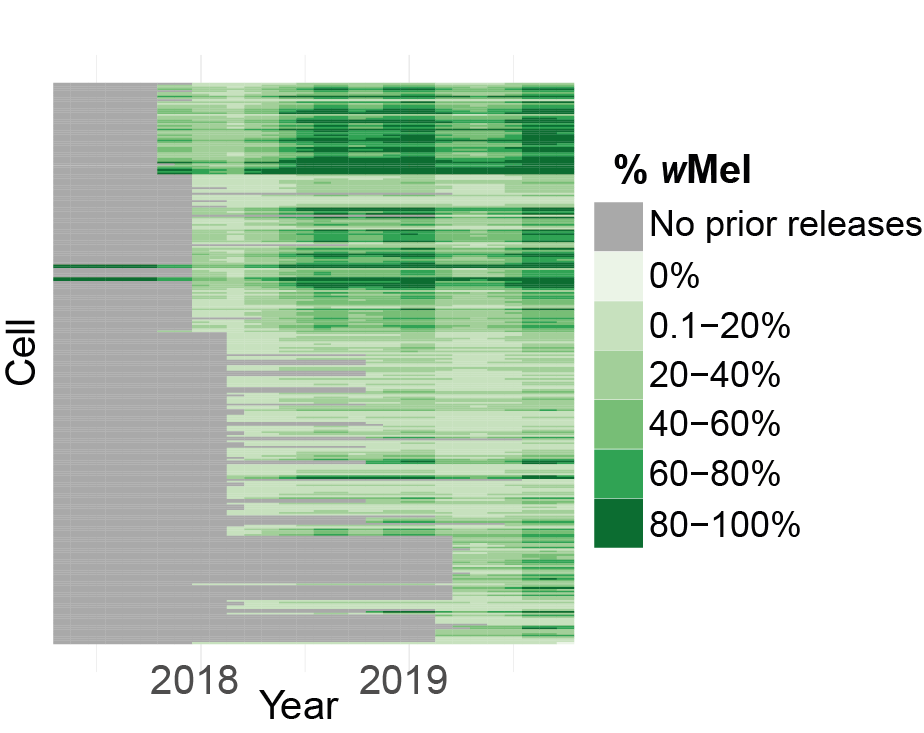

**Figure S1**  Values inferred by the %*w*Mel spatial model used to fill the gaps in Figure 1B.

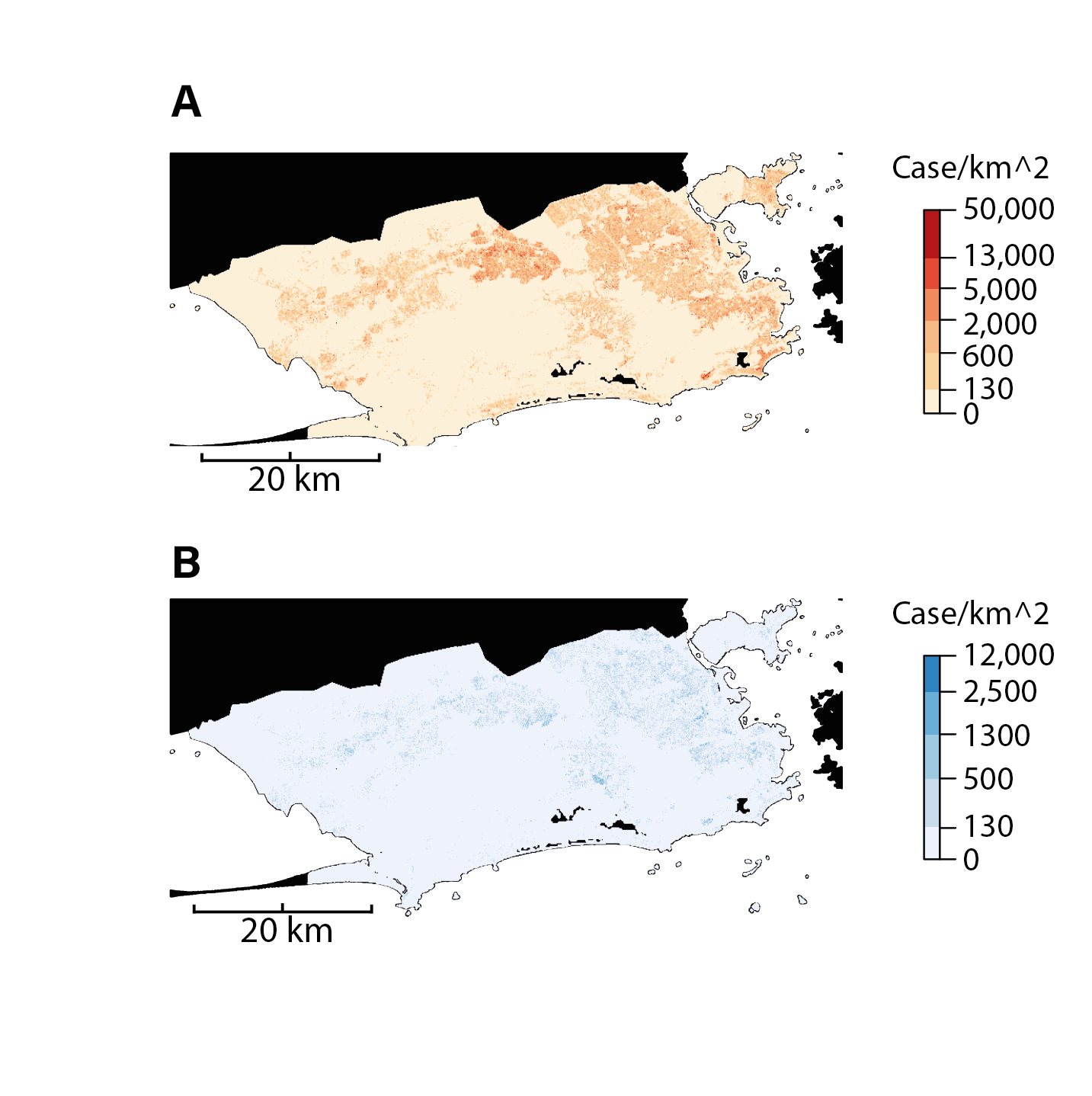

### **Figure S2** Map of total reported cases of **(A)** dengue and **(B)** chikungunya across the city.

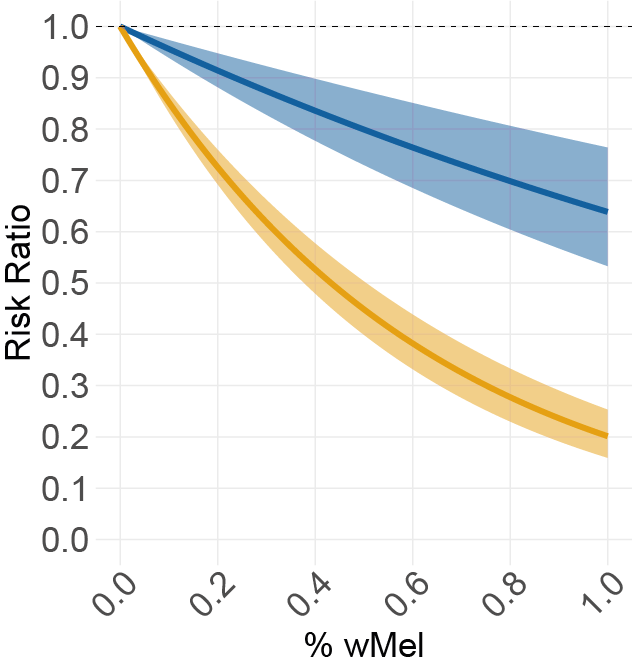

**Figure S3 :** Estimated reduction in incidence for dengue (orange) and chikungunya (blue) in places with a given amount of %*w*Mel relative to places where no *w*Mel has been detected. Model fitted with %*w*Mel as a continuous variable.

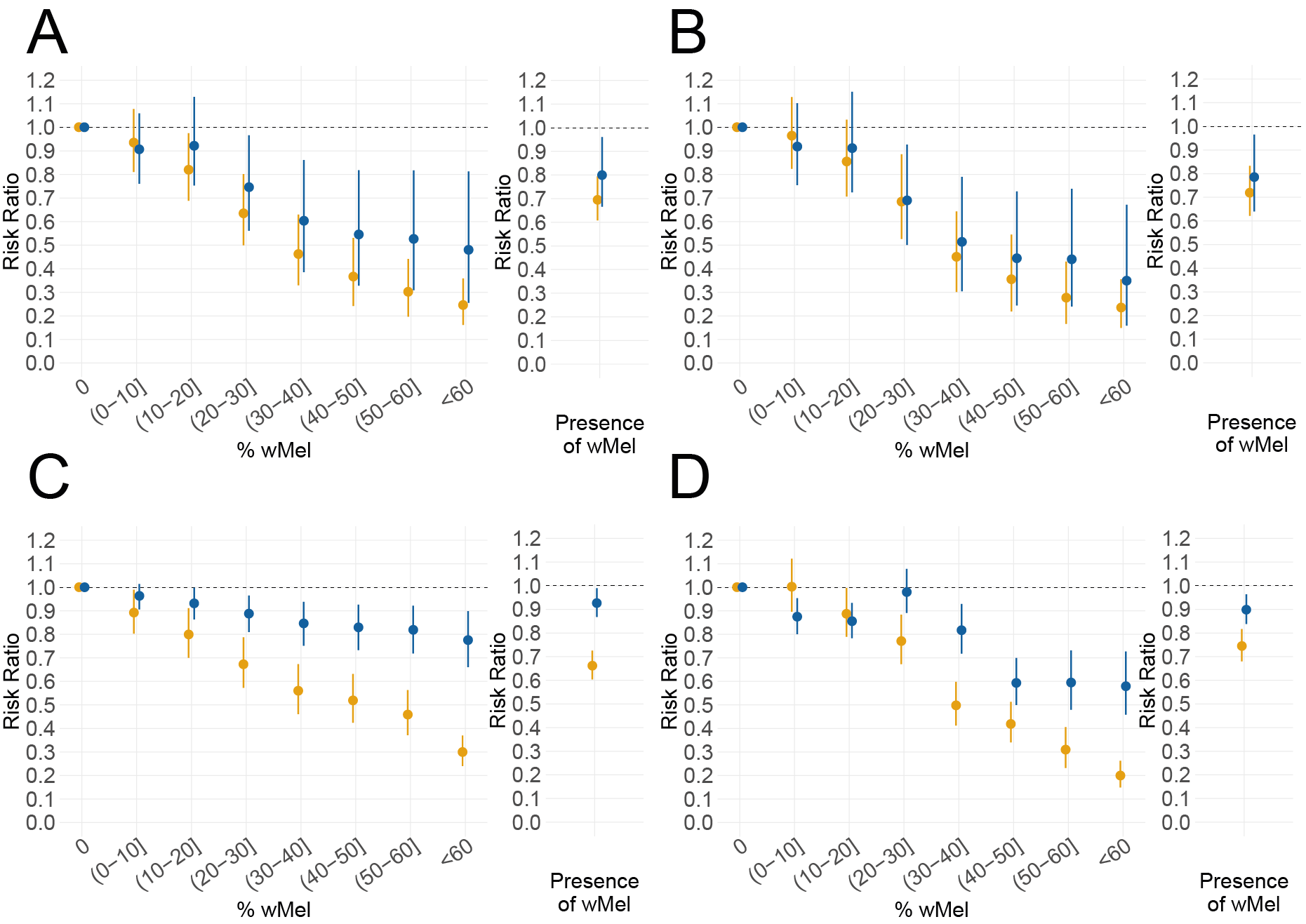

**Figure S4 Sensitivity analysis.** Models excluding space-time units where mosquitoes where released during the month (A) and additionally excluding cells where releases occurred in the previous month (B). Models with missing data completed by predicted values from mosquito count spatial model (C) and with all the data replaced by predicted values (D). Mean and 95% CI of the *w*Mel risk ratios given by the posterior distributions inferred with the model for all covariates for dengue (orange) and chikungunya (blue).

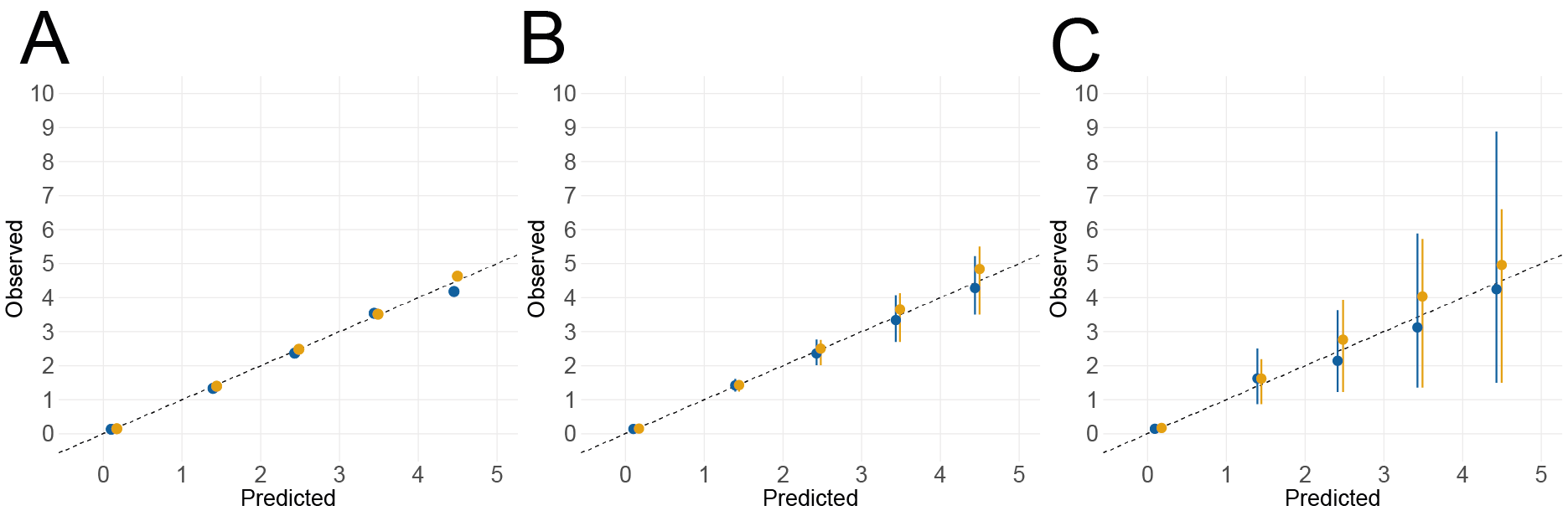

### **Figure S5** Precision plots giving the average observed values for dengue (orange) and chikungunya (blue). Uncertainty is given as the estimation of the 95% CI from a sample of different ways to split the dataset into training and testing. **(A)** Model fitting and predicting on the totality of the dataset **(B)** Model fitting on half of the dataset, prediction on the other half. Splitting is done randomly. **(C)** Model fitting on 80% of the dataset, prediction on the remaining 20%.Splitting is done by removing spatiotemporal chunks of dimensions 5km^2 * 1 year from the fitting set.
